# GlaucoRAG: A Retrieval-Augmented Large Language Model for Expert-Level Glaucoma Assessment

**DOI:** 10.1101/2025.07.03.25330805

**Authors:** Mohammad Aminan, S. Solomon Darnell, Mohammad Delsoz, Amin Nabavi, Claire Wright, Brian Jerkins, Siamak Yousefi

## Abstract

**Purpose:** Purpose: Accurate glaucoma assessment is challenging because of the complexity and chronic nature of the disease; therefore, there is a critical need for models that provide evidence-based, accurate assessment. The purpose of this study was to evaluate the capabilities of a glaucoma specialized Retrieval-Augmented Generation (RAG) framework (GlaucoRAG) that leverages a large language model (LLM) for diagnosing glaucoma and answering to glaucoma specific questions.

**Design:** Evaluation of diagnostic capabilities and knowledge of emerging technologies in glaucoma assessment.

**Participants:** Detailed case reports from 11 patients and 250 multiple choice questions from the Basic and Clinical Science Course (BCSC) Self-Assessment were used to test the LLM based GlaucoRAG. No human participants were involved.

**Methods:** We developed GlaucoRAG, a RAG framework leveraging GPT-4.5-PREVIEW integrated with the R2R platform for automated question answering in glaucoma. We created a glaucoma knowledge base comprising more than 1,800 peer-reviewed glaucoma articles, 15 guidelines and three glaucoma textbooks. The diagnostic performance was tested on case reports and multiple-choice questions. Model outputs were compared with the independent answers of three glaucoma specialists, DeepSeek-R1, and GPT-4.5-PREVIEW (without RAG). Quantitative performance was further assessed with the RAG Assessment (RAGAS) framework, reporting faithfulness, context precision, context recall, and answer relevancy.

**Main Outcome Measures:** The primary outcome measure was GlaucoRAG’s diagnostic accuracy on patient case reports and percentage of correct responses to the BCSC Self-Assessment glaucoma items, compared with the performance of glaucoma specialists and two benchmark LLMs. Secondary outcomes included RAGAS sub scores.

**Results:** GlaucoRAG achieved an accuracy of 81.8% on glaucoma case reports, compared with 72.7% for GPT-4.5-PREVIEW and 63.7% for DeepSeek-R1. On glaucoma BCSC Self-Assessment questions, GlaucoRAG achieved 91.2% accuracy (228 / 250), whereas GPT-4.5-PREVIEW and DeepSeek-R1 attained 84.4% (211 / 250) and 76.0% (190 / 250), respectively. The RAGAS evaluation returned an answer relevancy of 91%, with 80% context recall, 70% faithfulness, and 59% context precision.

**Conclusions:** The glaucoma-specialized LLM, GlaucoRAG, showed encouraging performance in glaucoma assessment and may complement glaucoma research and clinical practice as well as question answering with glaucoma patients.

## Introduction

Artificial intelligence (AI) models have demonstrated high accuracy in detecting several ophthalmic diseases, including diabetic retinopathy and glaucoma, from various imaging modalities over the past few years[1, 2, 3, 4, 5, 6, 7, 8, 9]. More recently, large language models (LLMs) have shown promise in answering clinical queries[10, 11, 12, 13, 14]. While LLMs like GPT-4.5-PREVIEW hold promise for automated question answering, there is a risk of producing hallucinated answers without grounding[15, 16].

Retrieval-Augmented Generation (RAG), which combines external, domain-specific knowledge, has been identified as a valuable approach with the capacity to improve the factual accuracy of LLM outputs[16]. RAG combines dense document embeddings with an LLM to generate responses that are validated through the embedded knowledge and typically provide evidence and grounding that minimizes hallucinations[1]. Previous RAG systems have performed well in radiology and pathology questions[2]. However, RAG-based models in ophthalmology remain relatively understudied.16 A head-to-head comparison of ChatGPT versus ophthalmologists on exam questions revealed that a general purpose LLM still performs lower on knowledge relevant to ophthalmologists, highlighting the necessity of domain-specialized RAG systems[6].

In this study, we developed GlaucoRAG, an augmented-retrieval LLM-based system to assess glaucoma using a rigorous glaucoma-specific knowledge base to facilitate more evidence-based question answering in glaucoma.

## 1. Methods

### 1.1 Knowledge Base Sources and Pre-Processing

The implementation of GlaucoRAG involves data curation, ancillary open-source tools, and a popular state-of-the-art LLM. The knowledge base for GlaucoRAG was compiled from three sources: three authoritative glaucoma textbooks, American Academy of Ophthalmology Basic and Clinical Science Course BCSC Section 10, Shields’ Textbook of Glaucoma (7th ed.), and Glaucoma (2nd ed.); 1,859 peer-reviewed articles published between 2000 and March 2020. Articles were retrieved from Ophthalmology, Progress in Retinal and Eye Research, Ophthalmology Glaucoma, American Journal of Ophthalmology, Survey of Ophthalmology, and Journal of Glaucoma using the query term “glaucoma” in the title or abstract; and 15 guideline documents on assessment and management of glaucoma published by American Academy of Ophthalmology, European Glaucoma Society, and Royal College of Ophthalmologists.

A limited set of 250 multiple-choice questions, without answers or explanatory content, were input into LLMs solely to evaluate LLM models. This use aligns with the AAO’s permitted usage guidelines and is consistent with the principles of Fair Use under U.S. copyright law, given the non-commercial, transformative nature of the work. No AAO answers, explanations, or proprietary educational material were reproduced or distributed in the study.

### 1.2 GlaucoRAG Architecture

GlaucoRAG’s inference components are R2R[17], an open source agentic RAG system and GPT-4.5-PREVIEW[18], a state-of-the-art LLM. Figure 1 shows the architecture of the GlaucoRAG. The system is composed of two coupled subsystems, an offline knowledge-ingestion pipeline and an online inference pipeline. Both system pipelines are orchestrated by Docker-Compose which exposes a single REST/WS endpoint via the open-source R2R framework (v3.6.5). Source code for every component is available in the public repository (commit paper-v1).

**Figure 1.**
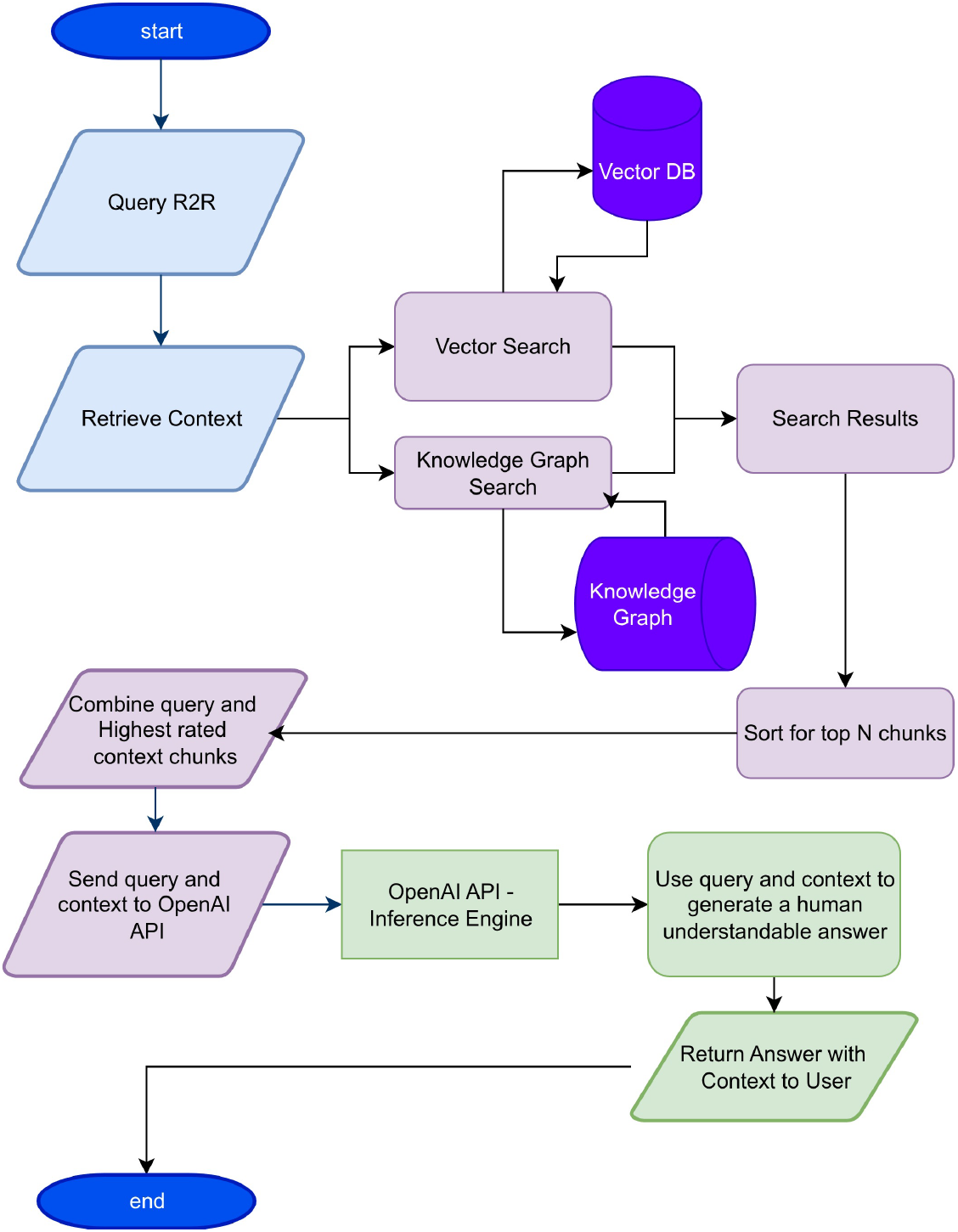
Architecture of the glaucoma specialized retrieval augmented generation (GlaucoRAG) framework.

#### 1.2.1 Offline knowledge ingestion pipeline

During ingestion, r2r_ingest.py iteratively traversed the corpus, eliminating duplicate titles, and segmented each document using an adaptive, progressive-fallback strategy. Initial segmentation employed 300-token chunks with 15-token overlap; if this precise mode failed, chunk size was reduced to 150 tokens with 10-token overlap, and, if necessary, a final fast mode ingested ≈ 1000-token chunks without overlap. Every resulting chunk was embedded with the default R2R dense encoder (OpenAI text-embedding-3-large) and stored in a Postgres-backed HNSW vector index using cosine similarity. Provenance was maintained by recording SHA-256 checksums for every processed file, while r2r_log-ging.py generated color-coded, timestamped logs that were written to /app/logs/ingest.log.

#### 1.2.2 Online inference pipeline

At query time, the retriever component is invoked through either r2r_chat_v4.py or r2r_query.py. R2R conducts an approximate nearest-neighbor search that initially returns 20 chunks; an optional cross-encoder re-ranking module (r2r_chat_v3_crossencoder.py) rescales similarity scores without introducing external text. The eight highest-ranked passages (≈ 3200 tokens total) are concatenated with the user prompt and forwarded to GPT-4.5-preview with deterministic generation settings (temperature 0; max_tokens = 1024). GlaucoRAG’s post-processing layer resolves numeric citations into human-readable article or textbook titles with page numbers, integrates them into the JSON response, and renders them inline in the chat interface. Robustness safeguards include an automatic retry that reduces context to four passages if the language model returns an empty string or malformed JSON; persistent failure then triggers an error message to the client.

### 1.3 Evaluation

Two evaluation tasks were used: (1) Case report task, a pilot set of 11 de-identified glaucoma case reports were drawn from EyeRounds, the open teaching repository of the University of Iowa’s Department of Ophthalmology and Visual Sciences[19]. Details of selection and redaction have been reported previously. Each case was presented to GlaucoRAG, two baseline LLMs (GPT-4.5-PREVIEW and DeepSeek-R1), and three fellowship-trained glaucoma specialists, who independently submitted a single best diagnosis. Reference diagnoses were taken from the EyeRounds key[19]. BCSC multiple-choice question task. Two hundred fifty glaucoma-focused items were extracted from the BCSC Self-Assessment bank, each consisting of a stem and four or five answer options. The questions were fed to the same three LLMs; responses were scored automatically against the official answer key. Each item was sent programmatically to GPT-4.5-PREVIEW (OpenAI Chat Completions, temperature 0, max_tokens 256) and DeepSeek-R1 (DeepSeek API v1, temperature 0, max_tokens 256) as a single-turn prompt containing only the question stem and answer options; the raw JSON responses were parsed automatically to extract the top-ranked answer choice. For both tasks, accuracy, expressed as the percentage of cases or questions answered correctly (top-1 match for the case reports).

Beyond conventional accuracy measurements, we utilized the RAG assessment (RAGAS) framework for a multi-faceted analysis. Four metrics were assessed by RAGAS: (1) Faithfulness, computed by extracting each claim in the answer and checking it against the cited passages; (2) Context Precision, the share of retrieved passages that contain ground-truth evidence; (3) Context Recall, the fraction of all ground-truth evidence found in the retrieved set; and (4) Answer Relevancy, a cross-encoder similarity score between the user’s question and the generated reply[20, 21].

All four metrics are reported on a 0 to 1, with higher values indicating better performance.

Accuracy was expressed as a binomial proportion (correct / total). Two-sided 95% confidence intervals (CI) were. Differences between independent proportions were assessed with a two-tailed two-proportion z-test. Statistical significance was defined as *p <* 0.05. All analyses were performed in Python 3.1.

## 2. Result

### 2.1 Case Report Tasks

On the case report task, GlaucoRAG correctly identified the reference diagnosis in 9 of 11, accuracy of 81.8% (95% CI, 52.3–94.9%). This exceeded the scores of standalone GPT-4.5-PREVIEW (8*/*11, 72.7%, 95% CI 43.4–90.3%) and DeepSeek-R1 (7 / 11, 63.6%, 95% CI 35.4–84.8%). The mean accuracy of the three fellowship-trained glaucoma specialists was ≈ 7/11 cases (66.7%). (Table 1)

**Table 1.** Accuracy of glaucoma experts, LLM Models, and GlaucoRAG at assessment of case reports, and BCSC self-assessment. LLM: Large language model; GlaucoRAG: Glaucoma specialized Retrieval-Augmented Generation; BCSC: Basic and Clinical Science Course; CI: Confidence interval.

| Task | Method | Correct / Total | Accuracy (95% CI) |
| --- | --- | --- | --- |
| Case reports | DeepSeek R1 (baseline) | 7 / 11 | 63.6% (35.4–84.8%) |
|  | GPT-4.5-PREVIEW | 8 / 11 | 72.7% (43.4–90.3%) |
|  | Glaucoma Attending 1 | 8 / 11 | 72.7% (43.4–90.3%) |
|  | Glaucoma Attending 2 | 7 / 11 | 63.6% (35.4–84.8%) |
|  | Glaucoma Attending 3 | 7 / 11 | 63.6% (35.4–84.8%) |
|  | GPT-4.5-PREVIEW + RAG (GlaucoRAG) | 9 / 11 | 81.8% (52.3–94.9%) |
| BCSC Self- Assessment | DeepSeek R1 (baseline) | 190 / 250 | 76.00% (70.3–80.9%) |
|  | GPT-4.5-PREVIEW | 211 / 250 | 84.40% (79.4–88.4%) |
|  | GPT-4.5-PREVIEW + RAG (GlaucoRAG) | 228 / 250 | 91.20% (87.0–94.1%) |

### 2.2 BCSC multiple-choice question task

Out of 250 glaucoma-focused items from the BCSC Self-Assessment bank, GlaucoRAG answered 228 correctly (91.2%, 95% CI 87.0–94.1%), versus 211 (84.4%, 95% CI 79.4–88.4%) for GPT-4.5-PREVIEW and 190 (76.0%, 95 %CI 70.3–80.9%) for DeepSeek-R1. A two-sample z-test confirmed that the 6.8-point margin over GPT-4.5-PREVIEW was statistically significant (p = 0.02), and the 15.2-point margin over DeepSeek-R1 was highly significant (*p <* 0.001). (Table 1)

### 2.3 RAGAS Quality Metrics

Table 2 shows details of the RAGAS quality metrics. RAGAS analysis on the 250 BCSC items yielded a faithfulness score of 0.70, context precision 0.59, context recall 0.80, and answer relevancy 0.91, indicating that most answers were well supported by retrieved evidence while maintaining high relevance despite a recall-oriented retrieval strategy.

**Table 2.** RAGAS evaluation results for GlaucoRAG framework. RAGAS: Retrieval-Augmented Generation Assessment; GlaucoRAG: Glaucoma Specialized Retrieval-Augmented Generation.

| Metrics | Values | Interpretation |
| --- | --- | --- |
| Faithfulness | 0.70 | Strong factual grounding with minimal hallucinations |
| Context Precision | 0.59 | Moderate retrieval specificity with optimization potential |
| Context Recall | 0.80 | Comprehensive knowledge coverage |
| Answer Relevancy | 0.91 | Excellent question-answer semantic alignment |

## 3. Discussion

In this study, we introduced and evaluated a RAG system, GlaucoRAG, designed to integrate expert curated textual knowledge for high accuracy question answering in glaucoma. Our results demonstrate a statistically significant improvement over non-RAG baselines. The performance boost observed with GlaucoRAG, 81.8% on case-based queries and 91.2% on BCSC multiple choice questions, exceeds the documented performance of non-RAG ophthalmic models. Consistent gains across small scale expert curated questions (18.2 percentage point improvement) and large-scale standardized assessments (6.8 percentage point improvement) validate the robustness of our approach.

The advent of LLM like GPT-4.5-PREVIEW has opened new horizons in ophthalmology, particularly in domains that require nuanced, evidence-based interpretation, such as glaucoma management. LLMs hold great promise for decision support, yet their utility in medicine is limited by well-known pitfalls, most notably the tendency to “hallucinate” clinically plausible but unverified statements, a problem rooted in generic training corpora, weak domain evaluation standards, and uneven data quality. Routing generation through a retrieval layer largely neutralizes this risk by obliging the model to anchor every statement in citable evidence, an approach that mirrors the way clinicians defend their conclusions. In addition, widespread adoption of LLMs in medicine still limited by privacy concerns. GlaucoRAG addresses these demands and demonstrating that a locally hosted, cost-effective system can deliver expert level accuracy while maintaining traceable, evidence-grounded responses. Similar to our model, Luo et. al. reported a retrieval augmented ophthalmic LLM aligning with expert consensus 84% of the time versus 46% without retrieval[16], and GPT4 alone achieved 48% accuracy on complex cases[22].

Comprehensive RAGAS evaluation revealed nuanced system performance characteristics beyond traditional accuracy metrics. The model achieved a faithfulness score of 70% indicates that seven out of ten generated responses matched retrieved source content factually, providing significant protection against hallucinations, a key requirement for clinical applications. Such a score places GlaucoRAG firmly within the clinical RAG space, where faithfulness values 65% are deemed acceptable. Notably, the relatively low context precision (59%) alongside high context recall (80%) shows that our retrieval policy prioritizes broad knowledge coverage over stringent precision filtering. This conservative approach minimizes information loss but increases contextual noise. Nevertheless, an 80% recall confirms that our knowledge base supplies the information needed to answer four of five clinical queries. In terms of relevancy, the score was 91% that closely supports the overall accuracy of 91.2% demonstrating robust question comprehension and appropriate response generation.

Despite these promising capabilities, several limitations merit discussion. First, our current implementation does not fuse ophthalmic imaging with textual data, an essential step for truly robust AI support in glaucoma, where accurate image interpretation is pivotal. As ophthalmic care is inherently multi-modal, future work should develop multi-modal RAG frameworks capable of retrieving and reasoning over both textual and imaging data. However, current LLMs have limited native image understanding capability. Second, although RAG mitigates the static knowledge cutoff of LLMs by injecting external documents, performance remains sensitive to retrieval quality; critical evidence may be missed or ranked low, reducing answer fidelity[15, 16]. Dynamic, continuously updated knowledge bases will be essential for near real-time clinical relevance.

In conclusion, we have demonstrated that a RAG approach significantly enhances automated glaucoma question answering across expert curated and standardized evaluations. The GPT4.5 + R2R architecture offers a promising, scalable model for evidence grounded medical question and answer.

## Data Availability

All data produced in the present study are available upon reasonable request to the authors, and will be made available online in a github repository.

https://store.aao.org/basic-and-clinical-science-course-section-10-glaucoma.html

https://webeye.ophth.uiowa.edu/eyeforum/glaucoma_cases.htm#gsc.tab=0

## Acknowledgments

This work was supported by NIH Grant R01EY033005 (SY). The funders had no role in study design, data collection and analysis, decision to publish, or preparation of the manuscript.

## Notes

### Competing Interest Statement

The authors have declared no competing interest.

### Author Declarations

We used 11 patient case reports and 250 questions from the Academy's Basic and Clinical Science Course (BCSC) on Glaucoma. The 11 glaucoma case reports are a subset from Eyerounds by https://webeye.ophth.uiowa.edu/eyeforum/glaucoma_cases.htm#gsc.tab=0. BCSC questions and answers can be acquired from https://store.aao.org/basic-and-clinical-science-course-section-10-glaucoma.html.

